# LITERATURE REVIEW THE EFFECT OF MINDFULNESS INTERVENTIONS ON BLOOD PRESSURE AND STRESS IN HYPERTENSION PATIENTS

**DOI:** 10.1101/2023.12.06.23299631

**Authors:** Adin Mu’afiro, Moses Glorino Rumambo Pandin, Nursalam

## Abstract

**Background:** Hypertension is a significant contributor to both illness and death on a global scale. One method of managing blood pressure is through adopting a healthy lifestyle and implementing blood pressure control measures, with or without the use of medication. The objective of the review paper is to elucidate the impact of mindfulness therapies on both blood pressure and stress levels in individuals diagnosed with hypertension.

**Method:** The research approach employed is literature observation, wherein relevant papers published between January 2019 and December 2023 are sought by conducting searches in four databases: Scopus and Science Direct. The search criteria include publications written in English, available in open access, and with full text. The study investigates the impact of mindfulness therapies on blood pressure and stress levels in individuals with hypertension, using Proquest and CINAHL as research databases

**Results:** A total of 14 papers have undergone evaluation. The primary emphasis lies in mindfulness techniques aimed at lowering blood pressure and alleviating stress. A mindfulness intervention was administered in group settings, with a duration of 2 hours per week for a period of 8 weeks. Subsequently, participants engage in physical activity inside the confines of their own homes, dedicating a minimum of 45 minutes per day for a duration of 6 to 10 days per week. They also monitor their blood pressure and stress levels. Blood pressure measurements are derived from the mean of three measurements done thrice day. Following the implementation of the mindfulness intervention, there was a notable reduction in both systolic and diastolic blood pressure among the intervention group, as compared to the first measurement. The investigation revealed substantial disparities between the experimental and control groups in the mean scores of positive stress (P = 0.001) and negative stress (P = 0.001).

**Conclusion:** Mindfulness can be a straightforward and cost-effective method for enhancing blood pressure and reducing stress.

## BACKGROUND

Hypertension is a prevalent non-communicable condition frequently observed in society. Hypertension, a leading contributor to illness and death, impacts around 1.2 billion individuals globally (Alhabeeb et al., 2023)(Katherine T Mills, Andrei Stefanescu, 2020). As per the World Health Organization (2011), around one billion individuals worldwide are afflicted with hypertension. There will be a significant rise in the occurrence of hypertension, with an estimated 29% of adults globally projected to be affected by 2025. Approximately 8 million individuals succumb to mortality annually as a consequence of hypertension (Katherine T Mills, Andrei Stefanescu, 2020).

According to the Indonesian Ministry of Health, the prevalence of hypertension in Indonesia is 33%, and this rate is consistently rising each year (Kemenkes RI, 2023). The data obtained from the regional health study (Riskesdas) conducted in 2018 indicates a rise in the prevalence of hypertension from 25.8% in 2013 (Riskesdas 2013) to 34.1% in 2018 (Kemenkes, 2020). In 2018, the prevalence of hypertension in Indonesia was 34.1%. The highest prevalence was observed in South Kalimantan at 44.3%, while the lowest was in Papua Province at 22.2% (Kemenkes RI, 2023).

Hypertension is commonly known as the silent killer due to the absence of symptoms in those with elevated blood pressure. The majority of patients report feeling well and free from illness. This condition significantly elevates the likelihood of developing heart disease, stroke, kidney failure, and other fatal conditions that entail substantial healthcare expenses (Kemenkes RI, 2023)

Environmental and behavioral factors are the primary contributors to a decline in blood pressure (Kohl-Heckl et al., 2022). One method of managing hypertension in patients is by the adoption of a healthy lifestyle and the implementation of disease control measures, with or without the use of medication (Kemenkes RI, 2023). Hypertension is intricately linked to lifestyle, mental well-being, and overall quality of life. If not well managed, it can significantly diminish one’s quality of life. In addition, stress has an impact on the quality of life of individuals with hypertension (Fisher et al., 2008). This leads to heightened emotional and behavioral alterations and interferes with cognitive and biological processes. The contemporary modifications in lifestyle contribute to a multitude of mental health issues. According to the World Health Organization (WHO), around 250 million individuals are impacted by mental disease (Babak, Motamedi, Mousavi, & Ghasemi, 2022).

Stress is a prevalent aspect of human existence, affecting those with hypertension as well. Any internal or external stimulation or stimulus that disturbs the balance of internal conditions will trigger a stress reaction and subsequent adjustment. According to Roy, a stimulus will influence the body’s coping processes through regulators and cognators (Brianna Chu; Komal Marwaha; Terrence Sanvictores; Derek Ayers, 2022) Nurses can facilitate and endorse measures that address the health and social requirements of the community, particularly among hypertensive individuals who encounter stress, through the use of mindfulness treatments (ICN, 2021).

Despite receiving sufficient medication, hypertension patients continue to exhibit suboptimal blood pressure levels, as indicated by multiple research findings. Effective blood pressure management is attained by less than 25% and 10% of individuals with hypertension in industrialized and underdeveloped nations, respectively (Lee et al., 2020) . Hence, there is a want for supplementary interventions aimed at diminishing blood pressure and stress levels

Prior studies have extensively examined the efficacy of mindfulness interventions in reducing stress and blood pressure (Babak, Motamedi, Mousavi, & Darestani, 2022; Eunjoo An Michael R. Irwin Lynn V. Doering Mary-Lyn Brecht Karol E. Watson Elizabeth Corwin Paul M. Macey, 2021; Loucks et al., 2023a; Moceri & Cox, 2019b; Noventi et al., 2022; Polcari et al., 2022; Ponte Márquez et al., 2019; ŞEN GÖKÇEİMAM et al., 2022; Torné-Ruiz et al., 2023; Vara-García et al., 2019; Wright et al., 2021). These studies have reported a significant reduction in blood pressure and stress levels among participants who underwent mindfulness interventions, as compared to those in the control group. The objective of this literature review is to present data on the efficacy of mindfulness therapies in lowering blood pressure and stress levels

## METHOD

### Strategy for searching and criteria for selecting

The search was conducted on pertinent publications published between January 2019 and December 2023, utilizing English, open access, and full-text options in four databases: Scopus and Science Direct. The search was conducted utilizing the PICO framework, which involved the population of individuals with hypertension, the intervention of attention and prayer, dhikr, or spirituality, comparison between intervention and control groups only, and the outcome of blood pressure and stress. Searches employ Boolean operators such as “AND” and ‘OR’ to combine the keywords “mindfulness” AND “stress” and “blood pressure” OR “hypertension”.

### Selection Studies

This literature evaluation was completed and presented following the PRISMA and Rayyan flowchart standards for selection studies. The next step was utilizing Rayyan to identify duplicate entries, followed by conducting abstract selection to ensure alignment with the predetermined inclusion and exclusion criteria. Subsequently, the investigation was conducted.

### Eligibility criteria

1. Individuals aged 18 years or older diagnosed with hypertension, regardless of whether they are receiving treatment.
2. Interventions: Mindfulness interventions, such as Mindfulness-Based Stress Reduction or similar approaches.
3. Comparator: The intervention group received mindfulness interventions, while the control group received standard treatment.
4. Results: The primary outcome measured was blood pressure and its variability. The research design options include randomized clinical trial, cross-sectional survey, prospective clinical trial, associative analytic, quasi-experimental, cluster randomized controlled trial, experimental study, two-group single-site clinical trial design, retrospective study, cross-sectional design, descriptive study, and a two-site randomized clinical trial.
5. Publication period: January 1, 2019 to December 1, 2023
6. Article type: Open access and full text

### Exclusion criteria

Hypertensive patients who are breastfeeding or pregnant and have previously undergone mindfulness therapy

## RESULTS

Results were obtained by considering the title, abstract, publication type, and language of the articles. Out of the remaining 38 articles, eligibility was assessed based on the full text. Ultimately, a total of 12 papers were included in the systematic observation

The author discovered a total of 1,326 publications across four databases: Scopus (128 articles), CINAHL (308 articles), Science Direct (284 articles), and Proquest (606 articles). Following the elimination of duplicate articles and the exclusion of papers that did not meet the PICO criteria, a total of 367 articles were included in the screening procedure. The article writing process involved utilizing the Rayyan tool to analyze the title and abstract. This yielded a total of 251 articles, with the remaining 116 articles specifically addressing the topic of mindfulness. Subsequently, an eligibility screening was conducted, taking into account the publication type and language, followed by an evaluation of acceptability using the complete text. A total of 29 papers were acquired for subsequent evaluation, with 14 articles meeting the inclusion criteria and 15 articles being discarded due to insufficient study quality.

Additional search results can be observed by referring to the literature search flow diagram depicted in Figure 1. The article provides the following information: Studies were identified through databases and registers. Before screening, duplicate records (n=8) were removed, as well as records marked as ineligible by automation tools (n=1268) and records removed for other reasons (n=50). Records were identified from various sources, including databases such as Scopus (n=128), CINAHL (n=308), Science Direct (n=284), and ProQuest (n=606). After identification, some records were excluded (n=251) and others were screened (n=367). Some reports were not retrieved (n=87) but were sought for retrieval (n=116). During screening, reports were excluded for various reasons, such as not being specific to hypertension (n=3), not discussing mindfulness (n=4), not discussing stress and blood pressure (n=4), or being an article review (n=4). A total of 29 reports were assessed for eligibility, and ultimately 14 studies were included in the review.

**Figure 1.**
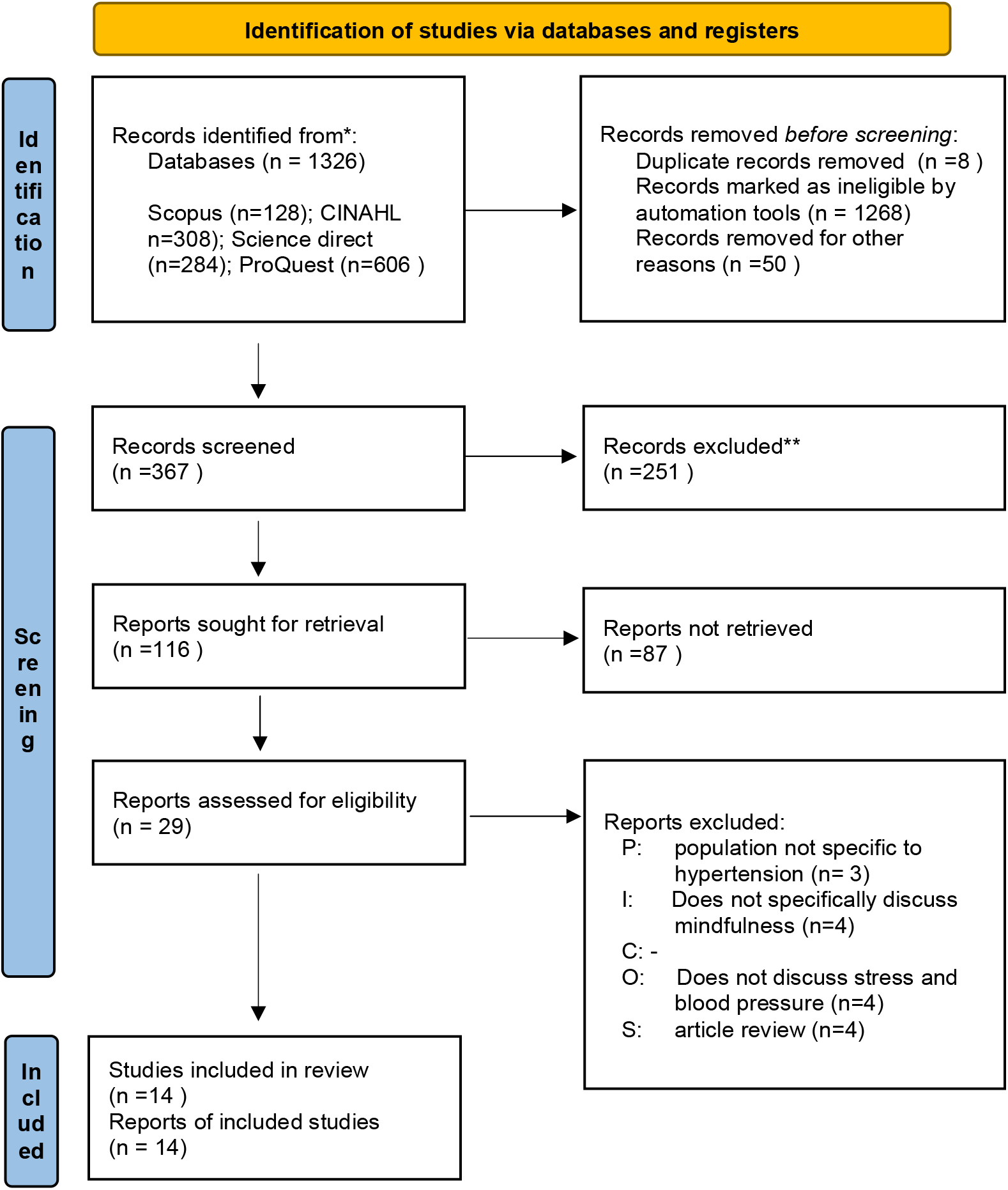
PRISMA flowchart.

The final results, namely 14 articles that are in accordance with the research, are as follows:

**Table 1.** Characteristics of reviewed studies.

| NO | Title, Author, Year | method | Results |
| --- | --- | --- | --- |
| 1 | Effects of Mindfulness-Based Stress Reduction on Blood Pressure, Mental Health, and Quality of Life in Hypertensive Adult Women: Randomized | **Design:** Randomized clinical trial<br>**Subjects:** 80 women with hypertension divided into 40 intervention groups and 40 control groups taken from the Imam Ali health care center in Iran | After administering the mindfulness intervention, the average systolic and diastolic blood pressure decreased significantly in the intervention group compared to the initial |
| | Clinical Pilot Study<br>(Babak, et.al, 2022) | **Variable:**<br>**Dependent:** blood pressure, stress, depression, anxiety, and quality of life<br>**Independent:** mindfulness-based stress reduction (MBSR)<br>**Instruments:**<br>Depression, Anxiety, and Stress Scale-21 (DASS-21) and 36-Item Short Form<br>Survey questionnaire (SF-36).<br>**Analysis:** independent t test, paired t test, and MANCOVA test. | value. control group systolic blood pressure ($140.18 \pm 14.27$ mmHg vs. $142.15 \pm 10.23$ mmHg and diastolic blood pressure $84.62 \pm 9.22$ vs. mmHg $88.51 \pm 8.54$ mmHg ( $P = 0.001$).<br>There was a significant increase in quality of life, stress, anxiety and depression scores in the intervention group ($P < 0.05$) |
| 2 | The Effects of Adapted Mindfulness Training on Participants With High Offices<br>Blood Pressure: MB-BP Study:<br>Randomized Clinical Trial<br>Source:<br>(Loucks et al., 2023a) | **Design :** randomized clinical trial (RCT)<br>**Subjects:** 201 hypertensive patients; 101 intervention groups and 100 control groups<br>**Variable:**<br>**Dependent:** Blood pressure and stress<br>**Independent:** Mindfulness training<br>**Instruments:**<br>Stress Scale;<br>Five-Faceted Mindfulness Questionnaire.<br>**Analysis**<br>t-test results | This study demonstrated a clinically significant reduction in systolic blood pressure in the mindfulness group compared to the control group ($n=100$).<br>The primary outcome was change in unmonitored clinic systolic blood pressure at 6 months.<br>showed a reduction in systolic blood pressure of 5.9 mm Hg (95% CI, $-9.1$ to $-2.8$ mm Hg) from baseline compared to the control group of 4.5 mm Hg at 6 months (95% CI, $-9.0$ to $-0.1$ mm Hg )<br>DASH (Dietary Approaches to Stop Hypertension) diet score (score 0.32 [95% CI, $-0.04$ to 0.67]), and mindfulness (score 7.3 [95%]) |
| 3 | Mindfulness-Based Exercise to Reduce Blood Pressure and Stress Priest<br>Source:<br>(Moceri & Cox, 2019a) | **Design:** Cross-section<br>**Subjects:** 11 priests,<br>**Variable:**<br>**Dependent:** Blood pressure, stress<br>**Independent:**<br>Mindfulness-Based Practice<br>**Instruments:**<br>Perceived Stress Scale (PSS), blood pressure observation sheet<br>**Analysis:**<br>t test; repeated measures analysis of variance (RM-ANOVA). | effectiveness of mindfulness-based interventions, demonstrating reductions in stress and blood pressure following implementation of mindfulness practices. 9.16 Average SBP of 121.06 mm Hg and DBP of 76.72 mm Hg meets the target blood pressure value below 150/80 mm Hg .<br>Mindfulness can lower blood pressure |
| 4. | Mindfulness-Based Meditation Sessions May Result in Reduction of Cardiovascular Risk in Hypertension Patients: A Pilot Study<br>(Trancoso Lopes et al., 2022) | **Design :**<br>Clinical trials<br>prospective,<br>Subjects:60 people<br>**Variable:**<br>**Dependent:** Blood Pressure<br>**Independent:** Single Mindfulness Based Meditation<br>**Instruments**<br>International Physical Activity Questionnaire (IPAQ-6)<br>**Analysis:**<br>one-way analysis of variance | There were no statistically significant differences between the treatment and control groups in hemodynamic variables. Morning systolic pressure was statistically lower after meditation SBP during the morning surge was statistically lower after the meditation session ($t=3.497$, $P < .05$). Additionally, mean blood pressure (MBP) values compared between sessions showed a significant trend after meditation, |
|  |  | (ANOVA) for ABPM data and two-way ANOVA for analyzing blood pressure | t= -2.648, P < .05 )<br>The results of our study showed a decrease in systolic blood pressure in the morning spike and downward trend in median blood pressure (MBP) values during the same day period (P = 0.057). However, clinical blood pressure did not change after MBSR. Therefore, one-time application of MBSR could potentially be an important tool to reduce cardiovascular risk in sedentary hypertensive women. |
| 5 | Benefits of Mindfulness Meditation in Lowering Blood Pressure and Stress in Arterial Hypertension Sufferers;<br>(Ponte Márquez et al., 2019) | **Design:** Randomized controlled trial<br>**Subjects:** 24 normal high BP patients and 18 grade I hypertension patients;<br>**Variable:**<br>**Dependent :** blood pressure, anxiety, stress and depression<br>**Independent:** mindfulness meditation<br>**Instruments :**<br>Perceived Stress Scale; Depression, Anxiety and Stress Scale (DASS-21); Five-Sided Mindfulness Questionnaire (FFMQ)<br>**Analysis:** Comparative descriptive | At baseline, the intervention group had nonsignificantly higher clinically measured blood pressure values, whereas the second group had similar ambulatory blood pressure monitoring (ABPM) values. At week 8, the intervention group had significantly lower ambulatory blood pressure monitoring scores than the control group (124/77 mmHg vs 126/80 mmHg (p < 0.05) and 108/65 mmHg vs 114/69 mmHg (p < 0.05) nighttime systolic blood pressure also had lower clinically measured systolic pressure values (130 mmHg vs 133 mmHg; p = 0.02). at week 8 the mindfulness group had lower clinically measured diastolic blood pressure values. |
| 6 | Effectiveness of Mindfulness-Based Group Therapy on Stress Perception, Cognitive Emotional Regulation, and Self-Care Behavior in Hypertension Sufferers ( Sabetfar et al., 2021) | **Design:** quasi-experimental design with pre-test, post-test, follow-up, and control group<br>**Subjects:** 32 candidates<br>**Variable:**<br>**Dependent:** Stress Perception, Cognitive Emotional Regulation, and Self-Care Behavior<br>**Independent:** Mindfulness-Based Stress<br>**Instruments :** Perceived Stress Scale<br>**Analysis:** repeated measures analysis of variance | The results showed significant differences between the experimental and control groups in the mean scores of positive stress (P=0.001), negative stress (P=0.001), positive emotions (P=0.001), negative emotions (P=0.001), treatment regimen (P=0.003), diet (P=0.011), and disease management (P=0.026) at post-test and follow-up. However, there was no significant difference between the food label mean scores (P=0.195). |
| 7 | Dhikr and Prayer Guidance Regarding Peace of Mind and Controlling Blood Pressure<br>( Samsualam , 2022) | **Design:** quasi-experimental with a pretest-posttest approach<br>**Subjects:** 24 respondents elderly people suffering from hypertension<br>**Variable:**<br>**Dependent:** Blood Pressure | The results of the study showed that the p value was 0.036 <0.05, meaning that it had no significant effect on the mental calm of elderly people with hypertension in the intervention group who |
|  |  | Control<br>**Independent** : Dhikr and Prayer for Peace of Mind<br>**Instrument** : observation sheet<br>**Analysis:** Wilcoxon Signed-Rank Test; t-test | received dhikr and prayer guidance. Blood pressure examination before and after in the intervention group obtained ap value of 0.000 <0.05, which means that there was an influence of the intervention group variable (systole) on blood pressure. |
| 8 | Effects of the Mindfulness-Based Blood Pressure Reduction (MB-BP) program on depression and neural structural connectivity<br>(Polcari et al., 2022) | **Design:** Randomized clinical trials (RCT)<br>**Subjects:** 36 people; 14 MBBP groups; 22 control groups<br>**Variable:**<br>**Dependent:** decrease in blood pressure<br>**Independent:** Mindfulness-Based Blood Pressure Reduction Program (MB-BP)<br>**Instruments:** American Heart Association guidelines<br>**Analysis:** Repeated measures ANOVA (RM ANOVA), standard linear regression. | Analysis of DTI data revealed significant group differences in several white matter neural tracts related to the limbic system and/or blood pressure. Specific changes in neural structural connectivity were significantly associated with measures of interoception and depression<br>It is concluded that MB-BP induces changes in brain structural connectivity that may mediate beneficial changes in depression and interoceptive awareness in individuals with hypertension. |
| 9 | Comparing the effectiveness of mindfulness-based stress reduction therapy and Islamic spirituality therapy on the quality of life of hypertensive heart patients (Shahbazi et al., 2022) | **Design:** experimental research, pretest-posttest design with a control group<br>**Subject:** 75 patients were selected and randomly divided into two experimental groups and one control group in such a way that each group consisted of 25 people.<br>**Variable:**<br>**Dependent:** Quality of life<br>**Independent** : MBSR spiritual therapy with an Islamic approach<br>**Instruments:** McNew quality of life questionnaire; John Kabbat's MBSR Protocol<br>**Analysis:** AVOVA and paired t test. | Finally, the overall quality of life score in the MBSR and spiritual therapy group improved significantly after the intervention ($P < 0.001$), but the overall quality of life score in the control group remained unchanged after the intervention ($P = 0.10$).<br>There was a significant difference between the three groups regarding the total quality of life score, so that the average difference score in the total quality of life score of the spiritual therapy group before and after the intervention was higher than the other groups. ($P < 0.001$) |
| 10 | Effects of mindfulness on lifestyle behavior and blood pressure: A randomized controlled trial ( Eunjoo An; et.al, 2021) | **Design:** a two-arm clinical trial at one site was designed<br>**Subjects:** 52 female participants, MAP (n = 20)<br>HPP (n = 16)<br>Controls (n=16)<br>**Variable:**<br>**Dependent:** Blood Pressure and Lifestyle behavior<br>**Independent:** mindfulness<br>**Instruments:** Rapid Eating and Activity Assessment for Patients (REAP) | The more significant reduction in blood pressure observed in the MAP group supports the evidence that the mindful meditation approach is a low-risk method for lowering blood pressure in adults with HTN.<br>There was a significant interaction between systolic blood pressure (SBP, $P = 0.005$) and diastolic blood pressure (DBP, $P = 0.003$) and the time difference between MAP and HPP. The |
| | | Exercise Behavior Questionnaire (EB) consisting of 6 items; Brief Treatment Questionnaire 1 (BMQ)<br>**Analysis:**<br>T-test. | mean decrease in SBP from baseline to week 13 for the MAP group was 19 mm Hg ($138 \pm 15$ mmHg-$119 \pm 6$ mm Hg) compared with 7 mmHg ($134 \pm 18$ mm Hg-$127 \pm 22$ mm Hg) in the HPP group .<br>Similarly, greater decreases in DBP were observed in the MAP group compared to the HPP group, 12 mm Hg ($89$ mm Hg ± $11$-$77 \pm 7$ mm Hg) and 1 mm Hg ($81 \pm 16$ mm Hg- $80 \pm 18$ mm Hg ), each.<br>Mean SBP was lower each week in the MAP group compared with the HPP group even though two subjects in the MAP group had stopped their antihypertensive medication. This trend was also observed in DBP, but the differences were smaller between the two groups. |
| 11 | Attention to stress and anxiety management in nursing students in clinical simulation: A quasi-experimental study<br>( Torné -Ruiz et al., 2023) | **Design :** Quasi-experimental a non-equivalent control group design<br>**Subjects :**<br>42 nursing students<br>**Variable:**<br>**Dependent:** blood pressure, heart rate, stress<br>**Independent:** mindfulness<br>**Instruments:**<br>VAS: self-administered stress analogue scale, STAI: State Trait Anxiety Inventory,<br>FFMQ: Five-Faced Mindfulness Questionnaire<br>**Analysis:**<br>Descriptive statistics and inferential statistics<br>Mann-Whitney and Wilcoxon tests | Although physiological characteristics in the experimental group (heart rate, $p = 0.048$, and diastolic blood pressure, $p = 0.032$) improved during the prebriefing phase, they were still statistically significantly lower. Anxiety ($p = 0.016$) and stress ($p = 0.029$) were also better controlled. During the debriefing session, both groups demonstrated statistically significant decreases in stress and anxiety as well as a number of physiological indicators. There were no discernible attentional shifts. |
| 12 | The Influence of Mindfulness Level on Drug Adherence in Hypertension sufferers<br>(Şen Gökçeimam et al., 2022) | **Design:** cross-sectional, descriptive study<br>**Subjects:** 68 hypertensive patients<br>**Variables:**<br>**Dependent:** drug adherence in Hypertension<br>**Independent :** minfulness<br>**Instruments;**<br>Socio-demographic data form and Mindful Attention Awareness scale (MAAS) and Modified Morisky Adherence scale (MMAS)<br>**Analysis:**<br>Chi-square, while the Student t-test or Mann-Whitney U test | There were no significant differences between groups in terms of adherence to medication when examined based on gender, education level, employment status and marital status.<br>In those with a family history of hypertension, low levels of medication adherence were significantly higher compared with moderate-high levels of medication adherence,<br>There was no statistically significant difference in medication adherence scores in |
|  |  | Multivariate logistic regression | terms of duration of hypertension. Mean MAAS scores were significantly higher in those with moderate-high medication adherence.<br>There was no statistically significant difference in medication adherence scores in terms of HT duration ($p=0.665$). Mean MAAS scores were significantly higher in those with moderate to high medication adherence ($p=0.004$) |
| 13 | The effectiveness of mindfulness-based stress reduction and similar vritti pranayama on reducing blood pressure, improving sleep quality and reducing stress levels in elderly people with hypertension<br>( Noventi et al., 2022) | **Design:** cross-sectional model<br>**Subject:** 30 people suffer from hypertension.<br>**Variables:**<br>**Dependent:** sleep quality, blood pressure, stress level<br>**Independent:** MBSR and samavitri pranayama programs.<br>**Instrument:** Indonesian version of the Pittsburgh Sleep Quality Index (PSQI); Mindfulness scale (Kentucky Inventory of Mindfulness Skills (KIMS) scale).<br>**Analysis:** Student's t test or chi square test | Overall, these findings show that the combination of MBSR and samavritti pranayama is successful in reducing blood pressure in elderly people with hypertension.<br>The mean systolic blood pressure in the intervention group showed a difference ($p, 0.001$). Mean systolic blood pressure before (155.00 mmHg) and after (130.00 mmHg). In the control group the average systolic blood pressure before (164.00 mmHg) and after (157.00 mmHg)<br>The mean diastolic blood pressure in the intervention group showed a difference ($p, 0.001$). Average diastolic blood pressure before (102.5.00 mmHg) and after (90.00 mmHg). In the control group the average diastolic blood pressure before (107.00 mmHg) and after (96.00 mmHg).<br>There was a difference in stress levels in elderly people with hypertension between the intervention group compared to the control group ($p=0.000$). The intervention group experienced a lower reduction in stress levels (7.5-2.2) than the control group (10.5-2.4).<br>The mediation program basically relieves mental and physiological complications in elderly people with hypertension. Specifically,<br>this research found that the combination of MBSR mediation and samavritti pranayama for 8 weeks succeeded in reducing hypertension, reducing anxiety scores and improving the quality of rest in the elderly. |
| 14 | Does Dispositional Mindfulness Predict Cardiovascular Reactivity to Emotional Stress in Prehypertension? Latent Growth Curve Analysis of the Serenity Study<br>Gabrielle<br>(Chin et al., 2021) | **Design:**<br>two-site randomized clinical trial<br>**Subject :** 153 participants,<br>**Variables:**<br>**Dependent:** blood pressure (BP) and heart rate (HR)<br>**Independent:** mindfulness<br>**Instruments:**<br>Self-report questionnaire; Five-Sided Mindfulness Questionnaire (FFMQ), Experience Questionnaire (EQ)<br>**Analysis :**<br>Assessment of suitability of the main model and LGCM; Familial alpha adjustment was applied to the effect of predictors of trait conscientiousness in each latent growth curve model ($p = 0.05$ split | Therefore, mindfulness-based interventions may be needed to address the benefits of mindfulness on stress physiology, as a biological mechanism thought to reduce cardiovascular risk and improve health.<br>For raw SPB reactivity, the mean intercept, or 'peak' reaction at time point two was 134.95 mmHg (SE = 0.90). The mean linear change was an increase of 1.95 mmHg per minute (SE = 0.35; $p < 0.001$), with a mean quadratic change of $-1.67$ mmHg (SE = 0.24; $p < 0.001$). Together, these latent growth parameters showed SBP increased significantly during the first three minutes of the anger recall task, then slowed and plateaued during the last three minutes of the stressor/<br>For raw DBP reactivity, the mean peak intercept, which occurred at the second time point, was 83.01 mmHg (SE = 0.69), with a mean linear change of 1.85 per minute (SE = 0.31; $p < 0.001$), and mean squared change $-1.08$ (SE = 0.21; $p < 0.001$), indicating similar reactivity curves for DBP during stressors. For raw HR reactivity, the mean peak intercept at the second time point was 73.87 bpm (SE = 0.91), with a mean linear change of 1.72 bpm (SE = 0.14; $p < 0.001$), and mean quadratic change was $-0.75$ (SE = 0.06; $p < 0.001$), indicating HR also increased in a quadratic fashion during the stressor. |

### Characteristics Study

The study examined the characteristics of different research designs used in the Randomized Control Trial (RCT). It included 1 articles that used a cluster randomized design (Rich et al., 2022); 4 articles that used quasi-experimental designs (Sabetfar et al., 2021; Samsualam, 2022; Shahbazi et al., 2022; Torné-Ruiz et al., 2023); and 3 articles that used cross-sectional designs (Moceri & Cox, 2019b; Noventi et al., 2022; Sen Gökçeimam et al., 2022). There is a single publication featuring a prospective clinical trial design authored by Trancoso Lopes et al. (Trancoso Lopes et al., 2022). The text mentions one article that describes a clinical trial conducted on a single group at one site (Eunjoo An; et.al, 2021) and another article that describes a randomized clinical trial conducted at two different sites (Chin et al., 2021).

In total, there were five randomized controlled trials (RCTs) conducted in the United States, including articles (Chin et al., 2021; Eunjoo et.al., 2021; Loucks et al., 2023a; Moceri & Cox, 2019b; Polcari et al., 2022). The studies were conducted in Spain by Márquez et al and Torné-Ruiz et al (Ponte Márquez et al., 2019; Torné-Ruiz et al., 2023), the articles were conducted in Iran by Babak, et.al. (2022) and Shahbazi et al. (2022) (Babak, Motamedi, Mousavi, & Darestani, 2022; Shahbazi et al., 2022)). In Indonesia, the articles were conducted by Noventi et.al. (2022) and Samsualam (2022); 1 One article was located in Brazil by Trancoso Lopes et al. (2022); and one article was conducted in Turkey by Şen Gökçeimam et al. (2022).

The five research enlisted volunteers from medical facilities and local populations. Every study included patient demographic information and baseline blood pressure measurements, which were evenly distributed between the treatment and control groups. All research participants were of diverse genders, with the exception of the studies conducted by Babak et al. (2022) and Trancoso Lopes et al. (2022), (Babak, et.al, 2022) and which only included female individuals. The participants’ age spanned from 45.8 to 60.3 years on average.

Among the 14 investigations conducted, one focused on clergy priest (Moceri & Cox, 2019a) another study featured students of nursing (Torné-Ruiz et al., 2023). and two studies examined elderly people with hypertension (Noventi et al., 2022; Samsualam, 2022).

### Participant characteristics

The research study included a total of 906 samples, with sample sizes ranging from 11 to 201. The study included individuals who were at least 18 years old and had hypertension, defined as having a systolic blood pressure of 180 mmHg or higher and/or a diastolic blood pressure of 80 mmHg or higher, with or without medication for hypertension. The average age in the intervention group was 48.67 ± 1.42 years, while in the control group it was 49.32 ± 2.31 years (P = 0.343) (Babak, Motamedi, Mousavi, & Darestani, 2022) with an average age of 45.8 ± 4.15 years (Trancoso Lopes et al., 2022).

### Intervention characteristics

The intervention aspects of the investigations were the use of mindfulness therapies, specifically Mindfulness-Based Stress Reduction (MBSR), which followed the theoretical framework proposed by John Kabbat Protocol (Shahbazi et al., 2022). A mindfulness intervention was administered for a duration of 2 hours per week over a period of 8 weeks, as reported by Wright et al. (2021) and Moceri & Cox (2019b). (Wright et al., 2021) (Moceri & Cox, 2019a) Additionally, a follow-up visit was conducted after 20 weeks, as documented by Babak, Motamedi, Mousavi, & Darestani (2022), Chin et al. (2021), Loucks et al. (2023a), Polcari et al. (2022), Ponte Márquez et al. (2019), and Sabetfar et al. (2021).

Participants had preliminary training, which included a collective introductory session, eight consecutive weekly group sessions of 2.5 hours each, and a total of 7.5 hours every group session per day. The suggested home mindfulness practice consists of a minimum of 45 minutes per day, for a total of 6 days each week. Psychiatric doctors (Ponte Márquez et al., 2019) offer training, whereas psychologists (Babak, et.al, 2022). conduct research.

A study conducted by Noventi et al. (2022) employed the MBSR and Samavittri Pranayama intervention regimens . Trancoso Lopes et al. (2022)found that mindfulness employs a method of expanding one’s awareness within a confined environment for a duration of 30 minutes. The Mindbody session comprises a concise elucidation of meditation and proper posture lasting 5 minutes, followed by a 15-minute body scan employing the body scan treatment approach. This is succeeded by a 5-minute instruction on breathing techniques, and concludes with a quick 5-minute talk to discuss the participants’ experience (Trancoso Lopes et al., 2022). Study conducted by Eunjoo An et al. (2021),

The researchers employed the Mindfulness Awareness Program (MAP) technique, which was created by the Mindfulness Awareness Research Center (MARC) at the University of California, Los Angeles (UCLA). MAP classes are instructed by trained educators. Every participant in the MAP program is provided with a practice guidebook and an audio recording containing instructions for conducting mindfulness sessions lasting 5 to 20 minutes. These resources are intended for personal use at home. Instructors of the Mindfulness and Acceptance Program (MAP) promote the regular practice of mindfulness, beginning with a duration of 5 minutes and gradually progressing to 20 minutes by the fifth week, as suggested by Wright et al. (2021). Physical activity is performed on a daily basis, first with a duration of 5 minutes and then progressing to 20 minutes by the fifth week (Eunjoo An; et.al, 2021).

Four investigations employed the Five Facet Mindfulness Questionnaire (FFMQ) as a measurement method (Chin et al., 2021; Loucks et al., 2023a; Ponte Márquez et al., 2019; Torné-Ruiz et al., 2023). A study employed the Kentucky Inventory of Mindfulness Skills (KIMS) scale, as developed by Noventi et al. in 2022. Vara-García et al. (2019) employed the Mindful Attention Awareness Scale (MAAS), while Şen Gökçeimam et al. (2022) utilized the Modified Morisky Compliance Scale (MMAS).., (2019) (Şen Gökçeimam et al., 2022).

### Blood pressure measuring instrument

All studies demonstrate the influence of mindfulness on both systolic and diastolic blood pressure. There are two techniques for evaluating blood pressure: clinic blood pressure and ambulatory blood pressure. Four studies conducted blood pressure assessments in clinical settings (Babak, et.al, 2022; Polcari et al., 2022; Ponte Márquez et al., 2019; Samsualam, 2022; Shahbazi et al., 2022). Two investigations employed clinical and ambulatory assessments to gather data (Ponte Márquez et al., 2019; Trancoso Lopes et al., 2022). The utilization of ambulatory blood pressure measures was employed in three studies (Chin et al., 2021; Eunjoo An Michael R. Irwin Lynn V. Doering Mary-Lyn Brecht Karol E. Watson Elizabeth Corwin Paul M. Macey, 2021; Loucks et al., 2023a).

Six investigations utilized Omron automated blood pressure monitors to evaluate blood pressure levels (Chin et al., 2021; Eunjoo An Michael R. Irwin Lynn V. Doering Mary-Lyn Brecht Karol E. Watson Elizabeth Corwin Paul M. Macey, 2021; Loucks et al., 2023b; Polcari et al., 2022; Ponte Márquez et al., 2019). There are no reports available for others. Prior to administering intervention, participants underwent training on the techniques for measuring blood pressure and conducting home monitoring. The participants recorded their blood pressure measurements in a book after a 5-minute break following the completion of the mindfulness exercise. These measurements were gathered weekly during the 6-week training session and again online 6 weeks later during the follow-up assessment (Eunjoo An; et.al, 2021).

### Systolic and diastolic blood pressure

Regarding systolic and diastolic blood pressure, a total of 14 papers demonstrated a decrease in blood pressure among the participants who underwent the mindfulness intervention. A study conducted by Loucks et al. (2023a) demonstrated a noteworthy decrease in systolic blood pressure among participants in the mindfulness group (n = 101) compared to those in the control group (n = 100). The unmonitored clinic systolic blood pressure decreased by 5.9 mm Hg (95% CI, −9.1 to −2.8 mm Hg) after 6 months, which was more than the control group’s decline of 4.5 mm Hg after 6 months (95% CI, −9.0 to −0.1 mm Hg).

Ponte Márquez et al., (2019) In the study conducted by Ponte Márquez et al. (2019) at the eighth week, the intervention group exhibited significantly lower scores in ambulatory blood pressure monitoring compared to the control group. The intervention group had blood pressure readings of 124/77 mmHg, while the control group had readings of 126/80 mmHg (p < 0.05). Additionally, the intervention group had readings of 108/65 mmHg, whereas the control group had readings of 114/69 mmHg (p < 0.05). Clinical measurements of nocturnal systolic blood pressure similarly showed reduced values for systolic pressure (130 mmHg vs. 133 mmHg; p = 0.02). There was a decrease in clinically determined diastolic blood pressure values among 8 mindfulness groups.

The study conducted by Babak, et.al. (2022), exclusively included female participants. It revealed a substantial decrease in both systolic and diastolic blood pressure among the intervention group as compared to the initial measurements. The control group had a systolic blood pressure of 140.18 ± 14.27 mmHg compared to 142.15 ± 10.23 mmHg, and a diastolic blood pressure of 84.62 ± 9.22 mmHg compared to 88.51 ± 8.54 mmHg (P = 0.001).

### Stress Measurement

Out of the 14 articles, 7 of them investigate the impact of mindfulness on stress. These articles include the works of Şen Gökçeimam et al. (2022), Loucks et al. (2023a), Moceri & Cox (2019b), Ponte Márquez et al. (2019), Sabetfar et al. (2021), Chin et al. (2021), and Noventi et al. (2022).

The Perceived Stress Scale (PSS) is the stress measurement tool used in several studies, including those conducted by Gu et al. (2023), Loucks et al. (2023a), Moceri & Cox (2019b), Noventi et al. (2022), Ponte Márquez et al. (2019), and Sabetfar et al. (2021) (Gu dkk., 2023; Loucks dkk., 2023a; Moceri & Cox, 2019b; Noventi dkk., 2022; Ponte Márquez dkk., 2019; Sabetfar dkk. al., 2021) .

Additionally, one article utilized a visual analog scale (VAS) to assess stress levels, as done by Torné-Ruiz et al. (2023) (Torné-Ruiz et al., 2023). The Depression, Anxiety, and Stress Scales (DASS-21) were employed in three articles (Babak, Motamedi, Mousavi, & Darestani, 2022; Ponte Márquez et al., 2019).

Eight articles have demonstrated the efficacy of mindfulness in reducing stress (Babak, Motamedi, Mousavi, & Darestani, 2022; Chin et al., 2021; Loucks et al., 2023a; Moceri & Cox, 2019a; Noventi et al., 2022; Ponte Márquez et al., 2019; Sabetfar et al., 2021; Torné-Ruiz et al., 2023).). Research findings indicate significant differences between experimental and control groups in terms of average scores for positive stress (P = 0.001) and negative stress (P = 0.001) (Sabetfar et al., 2021) (Moceri & Cox, 2019a).

There were differences in the stress levels of elderly hypertensive patients between the intervention group compared to the control group. The stress levels of elderly hypertension patients in the intervention group differed from those in the control group. The initial stress level, as indicated by the perceived stress scale (SPSS) score, in the intervention group was 11.3–3.1. However, following the intervention, it fell to 7.5–22 (p = 0.001). The control group had an initial score of 11.2-2.7, which decreased to 10.5-2.4 afterwards (p = 0.175)(Noventi et al., 2022).

## DISCUSSION

Hypertension stands as the primary determinant for mortality. Prevention entails altering modifiable risk factors. Hypertension prevention interventions, as recommended by the European Renal Association (ERA) and the International Society of Hypertension (ESH), involve following specific guidelines for managing arterial health. These guidelines include making lifestyle changes such as weight loss, reducing sodium intake, increasing potassium intake through food, engaging in regular physical activity and exercise, limiting alcohol consumption, quitting smoking, adopting a healthy diet, and improving stress management (Mancia et al., 2023).

Implementing a heart-healthy lifestyle is crucial for preventing the development of hypertensive heart disease, controlling elevated blood pressure levels, and reducing the risk of cardiovascular complications (Mancia et al., 2023).

Hypertension exhibits a strong correlation with lifestyle choices, mental well-being, and overall quality of life. If not effectively managed, it can lead to a range of issues, such as financial losses, reduced efficiency, and ultimately compromised well-being. Stress also impacts the quality of life of those with hypertension. Stress amplifies alterations in emotions and behaviors and interferes with cognitive and bodily processes (Fisher et al., 2008) (Babak, Motamedi, Mousavi, & Ghasemi, 2022).

According to the World Health Organization (WHO), around 250 million individuals are impacted by mental illness (Babak, Motamedi, Mousavi, & Ghasemi, 2022). Stress and worry have been linked to a higher likelihood of developing hypertension and experiencing cardiovascular events. Following the alleviation of a patient’s emotional distress, there may be a temporary surge in their blood pressure before it returns to its usual level. Exposure to severely traumatic life situations is also linked to an elevated risk. The European Renal Association (ERA) and the International Society of Hypertension (ESH) in 2023 established that the desired blood pressure goal for individuals with hypertension is a systolic pressure of 140 mmHg or a diastolic pressure of 90 mmHg (Mancia et al., 2023).

The findings of the literature study indicate that mindfulness-based stress release (MBSR) therapy has a substantial impact on reducing stress levels and altering mood, as well as lowering systolic and diastolic blood pressure (SBP and DBP). The Mindfulness Awareness Research Center (MARC) at UCLA defines mindfulness as “the process of actively and openly moment-by-moment observation of one’s physical, mental, and emotional experiences” (Eunjoo An; et.al, 2021). Mindfulness is very easy, cheap, and effective as an intervention to maintain blood pressure and reduce stress (Marino et al., 2021; Moceri & Cox, 2019a).

Environmental and behavioral factors are the greatest risk factors for increasing blood pressure. Therefore, the main intervention is to modify lifestyle and reduce stress among both through mindfulness (Kohl-Heckl et al., 2022).

Mindbody treatment approaches include mantra meditation, mindfulness meditation, spiritual meditation, guided imagery, progressive relaxation, yoga, tai chi, and qi gong (Kohl-Heckl et al., 2022). Individuals with hypertension most often used spiritual meditation (10.6%), yoga (5.7%), mindfulness meditation (3.2%), progressive relaxation (3.1%), mantra meditation (2.4%), guided imagery (1.9%), tai chi ( 1.5%), and qi gong (0.4%) (Kohl-Heckl et al., 2022)

The results of the literature review prove that the mindfulness intervention carried out for 45 minutes every day for 8 weeks was able to reduce systolic blood pressure and diastolic blood pressure and stress according to WHO targets, namely systolic blood pressure ≤ 130 mmHg and diastolic blood pressure ≤ 80 mmHg. Monitoring and measuring blood pressure at home is carried out three times a day in the morning, afternoon and evening.

## CONCLUSION

Interventions to reduce blood pressure and stress are very necessary for hypertensive patients to prevent complications. Mindfulness is an easy and cheap way to reduce blood pressure and stress for hypertension patients.

## Data Availability

All data produced in the present work are contained in the manuscript

## ACKNOWLEDGMENTS

Thank you to the Airlangga University lecturers and friends who have helped in writing this.

## REFERENCE

1. Alhabeeb, W., Tash, A. A., Alshamiri, M., Arafa, M., Balghith, M. A., Almasood, A., Eltayeb, A., Elghetany, H., Hassan, T., & Alshemmari, O. (2023). National Heart Center/Saudi Heart Association 2023 Guidelines on the Management of Hypertension. Journal of the Saudi Heart Association, 35(1), 16–39. 10.37616/2212-5043.1328

2. Babak, A., Motamedi, N., Mousavi, S. Z., & Darestani, N. G. (2022). Effects of Mindfulness-Based Stress Reduction on Blood Pressure, Mental Health, and Quality of Life in Hypertensive Adult Women: A Randomized Clinical Trial Study. Journal of Tehran University Heart Center, 17(3), 127–133.

3. Babak, A., Motamedi, N., Mousavi, S. Z., & Ghasemi, N. (2022). Effects of Mindfulness-Based Stress Reduction on Blood Pressure, Mental Health, and Quality of Life in Hypertensive Adult Women: A Randomized Clinical Trial Study. Journal of Tehran University Heart Center, 17(3), 127–133.

4. Brianna Chu; Komal Marwaha; Terrence Sanvictores; Derek Ayers. (2022). briana chu 2021 Physiology, Stress Reaction – StatPearls – NCBI Bookshelf (Jan 2023). StatPearls Publishing.

5. Chin, G. R., Greeson, J. M., Hughes, J. W., & Fresco, D. M. (2021). Does Dispositional Mindfulness Predict Cardiovascular Reactivity to Emotional Stress in Prehypertension? Latent Growth Curve Analyses from the Serenity Study. Mindfulness, 12(11), 2624–2634. 10.1007/s12671-021-01745-y

6. Eunjoo An Michael R. Irwin Lynn V. Doering Mary-Lyn Brecht Karol E. Watson Elizabeth Corwin Paul M. Macey. (2021). Mindfulness effects on lifestyle behavior and blood pressure: A randomized controlled trial. Health Science Reports, 4(2), 1–10. 10.1002/hsr2.296

7. Katherine T Mills, Andrei Stefanescu, J. H. (2020). The global epidemiology of hypertension. Nat Rev Nephrol ., 16(4), 223–237.

8. Kemenkes. (2020). Permenkes RI Nomor 21 Tahun 2020. Kementerian Kesehatan RI, 9(May), 6. https://www.slideshare.net/maryamkazemi3/stability-ofcolloids https://barnard.edu/sites/default/files/inline/student_user_guide_for_spss.pdf http://www.ibm.com/support http://www.spss.com/sites/dmbook/legacy/ProgDataMgmt_SPSS17.pdf https://www.n

9. Kemenkes RI. (2023). *Kemenkes RI*. https://sehatnegeriku.kemkes.go.id/baca/rilis-media/20230607/0843182/hipertensi-disebut-sebagai-silent-killer-menkes-budi-imbau-rutin-cek-tekanan-darah/

10. Kohl-Heckl, W. K., Schröter, M., & Cramer, H. (2022). Complementary medicine use in US adults with hypertension: A nationally representative survey. Complementary Therapies in Medicine, 65, 102812. 10.1016/j.ctim.2022.102812

11. Lee, E. K. P., Yeung, N. C. Y., Xu, Z., Zhang, D., Yu, C.-P., & Wong, S. Y. S. (2020). Effect and Acceptability of Mindfulness-Based Stress Reduction Program on Patients with Elevated Blood Pressure or Hypertension: A Meta-Analysis of Randomized Controlled Trials. Hypertension, 76(6), 1992–2001. 10.1161/HYPERTENSIONAHA.120.16160

12. Loucks, E. B., Schuman-Olivier, Z., Saadeh, F. B., Scarpaci, M. M., Nardi, W. R., Proulx, J. A., Gutman, R., King, J., Britton, W. B., & Kronish, I. M. (2023a). Effect of Adapted Mindfulness Training in Participants With Elevated Office Blood Pressure: The MB-BP Study: A Randomized Clinical Trial. Journal of the American Heart Association, 12(11). 10.1161/JAHA.122.028712

13. Mancia, G., Kreutz, R., Brunström, M., Burnier, M., Grassi, G., Januszewicz, A., Muiesan, M. L., Tsioufis, K., Agabiti-Rosei, E., Algharably, E. A. E., Azizi, M., Benetos, A., Borghi, C., Hitij, J. B., Cifkova, R., Coca, A., Cornelissen, V., Cruickshank, K., Cunha, P. G., … Kjeldsen, S. E. (2023). 2023 ESH Guidelines for the management of arterial hypertension The Task Force for the management of arterial hypertension of the European Society of Hypertension Endorsed by the European Renal Association (ERA) and the International Society of Hypertensi. *Journal of Hypertension*, Publish Ah(May), 1–198. 10.1097/hjh.0000000000003480

14. Marino, F., Failla, C., Carrozza, C., Ciminata, M., Chilà, P., Minutoli, R., Genovese, S., Puglisi, A., Arnao, A. A., Tartarisco, G., Corpina, F., Gangemi, S., Ruta, L., Cerasa, A., Vagni, D., & Pioggia, G. (2021). Mindfulness-based interventions for physical and psychological wellbeing in cardiovascular diseases: A systematic review and meta-analysis. Brain Sciences, 11(6). 10.3390/brainsci11060727

15. Moceri, J., & Cox, P. H. (2019a). Mindfulness-Based Practice to Reduce Blood Pressure and Stress in Priests. Journal for Nurse Practitioners, 15(6), e115–e117. 10.1016/j.nurpra.2019.01.001

16. Noventi, I., Sholihah, U., Nurhasina, S., & Wijayanti, L. (2022). The effectiveness of mindfulness based stress reduction and sama vritti pranayama on reducing blood pressure, improving sleep quality and reducing stress levels in the elderly with hypertension. Bali Medical Journal, 11(1), 302–305. 10.15562/bmj.v11i1.3108

17. Polcari, J. J., Cali, R. J., Nephew, B. C., Lu, S., Rashkovskii, M., Wu, J., Saadeh, F., Loucks, E., & King, J. A. (2022). Effects of the Mindfulness-Based Blood Pressure Reduction (MB-BP) program on depression and neural structural connectivity. Journal of Affective Disorders, 311, 31–39. 10.1016/j.jad.2022.05.059

18. Ponte Márquez, P. H., Feliu-Soler, A., Solé-Villa, M. J., Matas-Pericas, L., Filella-Agullo, D., Ruiz-Herrerias, M., Soler-Ribaudi, J., Roca-Cusachs Coll, A., & Arroyo-Díaz, J. A. (2019). Benefits of mindfulness meditation in reducing blood pressure and stress in patients with arterial hypertension. Journal of Human Hypertension, 33(3), 237–247. 10.1038/s41371-018-0130-6

19. Rich, T., Chrisinger, B. W., Kaimal, R., Winter, S. J., Hedlin, H., Yan, M., Zhao, X., Zhu, S., San-Lin, Y., Chien-An, S., Lin, J.-T., Hsing, A. W., & Heaney, C. (2022). Contemplative Practices Behavior Is Positively Associated with Well-Being in Three Global Multi-Regional Stanford WELL for Life Cohorts. International Journal of Environmental Research and Public Health, 19(20), 13485. 10.3390/ijerph192013485

20. Sabetfar, N., Meschi, F., & Taghvaei, M. H. (2021). Effectiveness of Mindfulness-based Group Therapy and Acceptance and Commitment on Perceived Stress, Emotional Cognitive Regulation and Self-Care Behaviors in Patients with Hypertension. JSabetfar, N., Meschi, F., & Taghvaei, M. H. (2021). Effectiveness of Mindfulness-Based Group Therapy and Acceptance and Commitment on Perceived Stress, Emotional Cognitive Regulation and Self-Care Behaviors in Patients with Hypertension. Journal of Criti, 14(4), 8–19.

21. Samsualam, M. (2022). Dhikr and Prayer Guidance on Peace of Mind and Blood Pressure Control. Indian Journal of Forensic Medicine & Toxicology, 16(2), 351–357. 10.37506/ijfmt.v16i2.17988

22. Şen Gökçeimam, P., Aydin Sünbül, E., Güçtekin, T., & Sünbül, M. (2022). The Effect of Mindfulness Level on Drug Adherence in Hypertension Patients. Namık Kemal Tıp Dergisi, 10(2), 212–218. 10.4274/nkmj.galenos.2022.96977

23. Shahbazi, K., Solati, K., Hosseinzadeh Taghvaei, M., Khaledifar, A., & Shah Nazari, M. (2022). Comparing the effectiveness of mindfulness-based stress reduction therapy and an Islam-based spirituality therapy on quality of life in hypertensive cardiac patients. Journal of Shahrekord University of Medical Sciences, 24(2), 47–53. 10.34172/jsums.2022.09

24. Torné-Ruiz, A., Reguant, M., & Roca, J. (2023). Mindfulness for stress and anxiety management in nursing students in a clinical simulation: A quasi-experimental study. Nurse Education in Practice, 66, 103533. 10.1016/j.nepr.2022.103533

25. Trancoso Lopes, B., Tanil Montrezol, F., Silva Terra, V. D., & Medeiros, A. (2022). A Single Mindfulness-Based Meditation Session Can Produce Reductions in Cardiovascular Risk in Hypertensive Patients: A Pilot Study. Alternative Therapies in Health & Medicine, 28(2), 18–23.

26. Vara-García, C., Romero-Moreno, R., Márquez-González, M., Mausbach, B. T., von Känel, R., Gallego-Alberto, L., Olmos, R., & Losada, A. (2019). Stress and Blood Pressure in Dementia Caregivers: The Moderator Role of Mindfulness. Clinical Gerontologist, 42(5), 512–520. 10.1080/07317115.2018.1554611

27. Wright, K. D., Klatt, M. D., Adams, I. R., Nguyen, C. M., Mion, L. C., Tan, A., Monroe, T. B., Rose, K. M., & Scharre, D. W. (2021). Mindfulness in Motion and Dietary Approaches to Stop Hypertension (DASH) in Hypertensive African Americans. Journal of the American Geriatrics Society, 69(3), 773–778. 10.1111/jgs.16947

